# Smoking Interaction with a Polygenic Risk Score for Reduced Lung Function

**DOI:** 10.1101/2021.03.26.21254415

**Authors:** Woori Kim, Matthew Moll, Dandi Qiao, Brian D. Hobbs, Nick Shrine, Phuwanat Sakornsakolpat, Martin D. Tobin, Frank Dudbridge, Louise V. Wain, Christine Ladd-Acosta, Nilanjan Chatterjee, Edwin K. Silverman, Michael H. Cho, Terri H. Beaty

**Author notes:** Correspondence: Terri H. Beaty, Ph.D., 615 N. Wolfe Street, Baltimore, MD 21205, and/or Michael H. Cho, M.D., M.P.H., 181 Longwood Avenue, Boston, MA 02115. These authors contributed equally (Woori Kim, Matthew Moll). Co-senior authors (Michael H. Cho, Terri H. Beaty).

## Abstract

**Importance:** Risk to airflow limitation and Chronic Obstructive Pulmonary Disease (COPD) is influenced by combinations of cigarette smoking and genetic susceptibility, yet it remains unclear whether gene-by-smoking interactions contribute to quantitative measures of lung function.

**Objective:** Determine whether smoking modifies the effect of a polygenic risk score’s (PRS’s) association with reduced lung function.

**Design:** United Kingdom (UK) Biobank prospective cohort study.

**Setting:** Population cohort.

**Participants:** UK citizens of European ancestry aged 40-69 years, with genetic and spirometry data passing quality control metrics.

**Exposures:** PRS, self-reported pack-years of smoking, ever-versus never-smoking status, and current-versus former-/never-smoking status.

**Main Outcomes and Measures:** Forced expiratory volume in 1 second (FEV_1_)/forced vital capacity (FVC). We tested for interactions with models including the main effects of PRS, different smoking variables, and their cross-product term(s). We also compared the effects of pack-years of smoking on FEV_1_/FVC for those in the highest versus lowest decile of predicted genetic risk for low lung function.

**Results:** We included 319,730 individuals (24,915 with moderate-to-severe COPD). The PRS and pack-years were significantly associated with lower FEV_1_/FVC, as was the interaction term (β [interaction] = −0.0028 [95% CI: −0.0029, −0.0026]; all p < 0.0001). A stepwise increment in estimated effect sizes for these interaction terms was observed per 10 pack-years of smoking exposure (all p < 0.0001). There was evidence of significant interaction between PRS with ever/never smoking status (β [interaction] = −0.0064 [95% CI: −0.0068, −0.0060]) and current/not-current smoking (β [interaction] = −0.0091 [95% CI: −0.0097, −0.0084]). For any given level of pack-years of smoking exposure, FEV_1_/FVC was significantly lower for individuals in the tenth compared to the first decile of genetic risk (p < 0.0001). For every 20 pack-years of smoking, those in the top compared to the bottom decile of genetic risk showed nearly a twofold reduction in FEV_1_/FVC.

**Conclusions and Relevance:** COPD is characterized by diminished lung function, and our analyses suggest there is substantial interaction between genome-wide PRS and smoking exposures. While smoking has negative effects on lung function across all genetic risk categories, effects of smoking are highest in those with higher predicted genetic risk.

**Key Points:** *Question:* Does cigarette smoking modify the magnitude of genetic effects on forced expiratory volume in 1 second (FEV_1_)/forced vital capacity (FVC)?

*Findings:* FEV_1_/FVC is influenced by PRS-by-smoking interactions. Smoking was detrimental across all categories of predicted genetic risk, though it was worse for those with highest predicted genetic risks. For every reported 20 pack-years of reported smoking, individuals in the top compared to the bottom decile of genetic risk showed nearly twice the reduction in FEV_1_/FVC.

*Meaning:* Elucidating mechanisms for the interaction between smoking and genetic risk could yield greater insight into the pathogenesis of chronic obstructive pulmonary disease (COPD).

## Introduction

Chronic obstructive pulmonary disease (COPD) is characterized by airflow obstruction, traditionally defined by a low forced expiratory volume in 1 second (FEV_1_)/forced vital capacity (FVC), and cigarette smoking is the greatest environmental risk factor ^1,2^. Only a minority of smokers develop COPD^3,4^, and genetic factors account for some of this variation in susceptibility, with ∼40% of the variability in spirometric measures of pulmonary function attributed to genetic variation ^5–7^. Therefore, it has long been thought that airflow obstruction may develop partially as the result of gene-by-smoking interactions.

Despite the important contribution of both smoking and genetic factors to lung function, compelling evidence for gene-by-smoking interactions has been limited. Genome-wide interaction studies have identified a handful of spirometric- and COPD-associated loci that appear to interact with smoking status,^8–14^ suggesting at least a portion of the variability in spirometric measures of lung function may be attributable to gene-by-smoking interactions. A major challenge in identifying gene-by-smoking interactions on lung function and risk to COPD is that individual genetic variants tend to be of small effect size and account for a low degree of phenotypic variability in lung function, diminishing the power to detect gene-by-smoking interactions.

Pooling individual GWAS variants into a single genetic risk score can account for a greater proportion of phenotypic variability^15–20^, and should improve power to detect interactions. Genetic risk scores have been used to investigate gene-by-environment interactions in psychiatric^21^ and cardiovascular diseases^22^. Aschard and colleagues^23^ were unable to detect individual single nucleotide polymorphism (SNP)-by-smoking interactions for FEV_1_/FVC for 26 variants identified as significant in a genome-wide joint meta-analysis of SNP-by-smoking associations of pulmonary function^14^; however, when the authors summed these variants to create a genetic risk score, they found evidence of interaction between the genetic risk score and ever smoking status^23^. By contrast, Shrine et al. performed the largest GWAS of lung function to date, developed a genetic risk score including estimated effects of 279 variants showing significant effects on lung function, and reported no evidence of interaction between this genetic risk score and ever smoking status ^19^, though the authors did observe an interaction of the genetic risk score with ever smoking status on moderate-to-severe COPD. We constructed a polygenic risk score (PRS) based on GWASs of FEV_1_ and FEV_1_/FVC^19^ that explained more of the variability in lung function than seen with the 279-variant risk score used by Shrine et al. (∼30% versus <10%) ^20^, and here we further investigate gene-by-smoking interactions.

We hypothesized that multiple measures of smoking exposure would significantly modify the effect of this genome-wide PRS on FEV_1_/FVC (i.e. because it is associated with lower lung function) in the United Kingdom (UK) Biobank population-based cohort.

## Methods

### Study population

We included participants from the UK Biobank, a cohort of over 500,000 individuals aged 40-69 years ^24^. All participants provided written informed consent and study protocols were approved by local institutional review boards/research ethics committees. Participants were excluded if spirometry or genetic data did not meet quality control standards; further details on the impact of these inclusion and exclusion criteria are shown in Figure S1. Quality control (QC) of spirometric data has been previously described ^18,19,24^. Briefly, to determine lung function, FEV_1_ and FVC were derived from the spirometry volume-time series data at the time of study enrollment, as previously reported^19^.

Genotyping was performed as previously described^19^, using Axiom UK BiLEVE array and Axiom Biobank array (Affymetrix, Santa Clara, California, USA) and imputed to the Haplotype Reference Consortium version 1.1 panel (accepting imputation accuracy R^2^ > 0.5). We dropped variants with minor allele frequency < 0.01 and those showing deviation from Hardy-Weinberg equilibrium (p < 1e-6). We used only subjects of European ancestry based on a combination of self-reported ethnicity and k-means clustering of principal components of genetic ancestry, as previously reported^19^.

### Statistical analyses

All analyses were done in R version 4.0.3 (www.r-project.org). Normality of continuous variables was assessed by visual inspection of histograms. Results are reported as mean ± standard deviation or median [interquartile range], as appropriate. Differences in continuous variables were assessed with Student t-tests or Wilcoxon tests, and categorical variables were compared by ANOVA or Kruskal-Wallis tests, as appropriate.

### Overview of study design

The primary outcome was the FEV_1_/FVC ratio, as clinical COPD is characterized by airflow obstruction (FEV_1_/FVC < 0.7), and severity graded based on decrements in FEV_1_ % predicted ^1,2^. We first assessed whether three measures of smoking exposure (see below) modified the effect of a PRS on quantitative measures of FEV_1_/FVC. We then considered the joint effects of smoking exposures and being in the highest versus lowest decile of the PRS (i.e. highest versus lowest categories of predicted genetic risk). We examined “norms of reaction” for the relationship between pack-years of smoking and FEV_1_/FVC for those in the tenth compared to the first decile of predicted genetic risk.

### Smoking exposures

We examined three measures of cigarette smoking exposure: 1) pack-years of smoking, 2) ever-versus never-smoking status, and 3) current smoker versus former-/never-smoking status. All smoking information was obtained by self-report. Pack-years of smoking was examined as continuous and categorical (pack-year categories: ≤10, 10.1-20, 20.1-30, 30.1-40, 40.1-50 and >50; where the reference group is ≤10 pack-years) variables. ‘Ever-smokers’ included individuals reporting current smoking, smoking most days, or smoking occasionally. ‘Never smokers’ included those who smoked less than 100 cigarettes in their lifetime. ‘Current smokers’ included those who reported current smoking, and ‘former smokers’ included non-current smokers who smoked more than 100 or more cigarettes in their lifetime.

### Polygenic risk score for lung function

A polygenic risk score (PRS) for lung function was calculated as previously described ^20^. Briefly, this PRS was based on genome-wide association results for FEV_1_ and FEV_1_/FVC in UK Biobank and SpiroMeta^19^, and was developed using a penalized regression framework accounting for linkage disequilibrium^25^. PRSs were calculated for FEV_1_ and FEV_1_/FVC, and then summed into a composite PRS, which was scaled and centered. The PRS was oriented such that a higher PRS was associated with lower FEV_1_ and FEV_1_/FVC.

### Interaction analyses

We performed multivariable linear regressions of FEV_1_/FVC on main effects of the combined PRS, smoking exposure, and cross-product interaction terms. We included covariates age, age^2^, sex, height, genotyping array and the first 10 principal components of genetic ancestry in the linear regression model. Age was scaled and centered prior to squaring. We also performed stratified analyses amongst those in the lowest and highest deciles of the PRS, separately for never and ever smokers.

Investigation of gene-by-environment interactions has been considered to be deviation from either an additive or multiplicative model. Therefore, we additionally examined the joint effects of smoking and PRS to assess a departure of the observed joint effect from the expected effect under an additive model. We focused on comparing the 1^st^ and 10^th^ deciles of PRS as previously done^20^. We created a categorical variable with mutually exclusive strata formed by the cross classification of smoking and PRS (10^th^ vs. 1^st^ decile). The reference category was the group with the lowest relative smoking exposure (e.g. pack-years ≤ 10) in the first PRS decile. We then constructed multivariable linear regression models to evaluate the effects of this categorical variable on FEV_1_/FVC. The expected effect for those in the highest decile with the highest smoking exposure was estimated under an additive model, and calculated by summing the estimated effect size for the lowest decile vs. the highest smoking exposure group and the highest decile vs. the lowest smoking exposure group.

### Norms of Reaction

A “norm of reaction” describes the relationship between a phenotype and environmental exposure for a given genotype^27^. We assessed norms of reaction for pack-years of smoking and FEV_1_/FVC for those in the lowest and highest deciles of predicted genetic risk. We plotted pack-years of smoking versus FEV_1_/FVC, stratifying by lowest and highest deciles of genetic risk. We then compared the slopes of these lines with an Analysis of Covariance (ANCOVA) using the rstatix R package^28^. For the purposes of clinical interpretability, we trained multivariable linear regression models to assess the effect of 20 pack-years of smoking on FEV_1_/FVC for those in the highest and lowest deciles of predicted genetic risk, adjusting for the covariates detailed as above. As sensitivity analyses, we repeated these analyses in ever smokers and in a dataset excluding all related individuals; to select unrelated individuals, we removed at least one individual from each related pair with kinship coefficient > 0.0625, favoring the inclusion of COPD cases. We also transformed reported pack-years of smoking (ln, scaling and centering, and rank normalization) to ensure that the effects of interaction terms were not due to misspecification of the main effects of smoking.

## Results

### Characteristics of study participants

Characteristics of study participants are shown in Table 1. We included 319,730 participants; 24,915 participants met criteria for moderate-to-severe COPD cases (Global Initiative for Chronic Lung Disease (GOLD) spirometry grades 2-4^1^), 38,713 had preserved ratio with impaired spirometry (PRISm)^29^, and 256,102 met criteria for GOLD spirometry grades 0/1.

**Table 1:** Characteristics of study participants. All participants were of European ancestry with high-quality genetic data and spirometry. Controls included those who met the spirometry criteria for Global Initiative for Chronic Obstructive Lung Disease (GOLD)^1^ spirometry grade 0/1, and cases included GOLD 2-4 participants (i.e. those with moderate-to-severe COPD). PRISm = preserved ratio with impaired spirometry, defined as FEV_1_/FVC > 0.7 with FEV_1_ % predicted < 80%^29^. P-values were calculated with ANOVA or Kruskal-Wallis tests across three categories for normal and non-normal variables, respectively.

|  | *Overall* |
| --- | --- |
| n | 319730 |
| Age (mean (SD)) | 56.45 (8.02) |
| Sex (No. (%) male) | 141864 (44.4) |
| Pack years of smoking (median [IQR]) | 0.00 [0.00, 11.00] |
| Former/Never smoker (No. (%)) | 287445 (89.9) |
| Current smoker (No. (%)) | 32242 (10.1) |
| Ever smoker (No (%)) | 146679 (45.9) |
| FEV1 % predicted (mean (SD)) | 92.17 (16.01) |
| FEV1/FVC (mean (SD)) | 0.76 (0.06) |
| Decile of polygenic risk score (%) |  |
| Lowest | 31973 (10.0) |
| Highest | 31973 (10.0) |

### Interaction of a polygenic risk score with smoking

The PRS was weakly correlated with pack-years of smoking (r = 0.041, p < 0.0001; Figure S2). The relationship between PRS and FEV_1_/FVC stratified by pack-years of smoking categories is illustrated in Figure 1. In multivariable analyses (Table 2), the main effects of the PRS (β=-0.0304 [95% CI: −0.0307,-0.0302]) and pack-years of smoking categories were associated with FEV_1_/FVC (all p<0.0001), and estimated effect sizes increased with each incremental category of smoking exposure. The same incremental trend for interaction terms between PRS and pack-years of smoking categories (all tests for interaction yielded p < 0.0001) were observed. Considering pack-years of smoking as a continuous variable (Table S1), the cross-product interaction term was also associated with FEV_1_/FVC (β [interaction]=-0.0028 [95% CI: −0.0029, −0.0026], p < 0.0001). We also performed three transformations of pack-years of smoking, and the PRS*pack-years interaction term was significant in each instance (all p < 0.0001, Table S2).

**Table 2:** Multivariable linear regression of the form FEV_1_/FVC ∼ PRS + Pack years of smoking + PRS*pack-years of smoking + covariates. Covariates include age, age^2^, sex, height, genotyping array, principal components of genetic ancestry. Pack years of smoking is included as a categorical variable with ≤ 10 pack-years as the reference group. PRS = polygenic risk score.

| *Variable* | *Beta (95% CI)* | *P* |
| --- | --- | --- |
| PRS | -0.03 (-0.031--0.03) | < 1e-256 |
| 11-20 pack years | -0.011 (-0.012--0.01) | 1.70E-225 |
| 21-30 pack years | -0.014 (-0.015--0.014) | < 1e-256 |
| 31-40 pack years | -0.026 (-0.027--0.025) | < 1e-256 |
| 41-50 pack years | -0.034 (-0.035--0.033) | < 1e-256 |
| >50 pack years | -0.04 (-0.041--0.038) | < 1e-256 |
| PRS X 11-20 pack years category | -0.0038 (-0.0046--0.0031) | 4.80E-24 |
| PRS X 21-30 pack years category | -0.0068 (-0.0077--0.006) | 1.60E-56 |
| PRS X 31-40 pack years category | -0.013 (-0.014--0.012) | 5.50E-133 |
| PRS X 41-50 pack years category | -0.015 (-0.017--0.014) | 1.90E-101 |
| PRS X >50 pack years category | -0.017 (-0.019--0.016) | 3.60E-134 |

**Figure 1:**
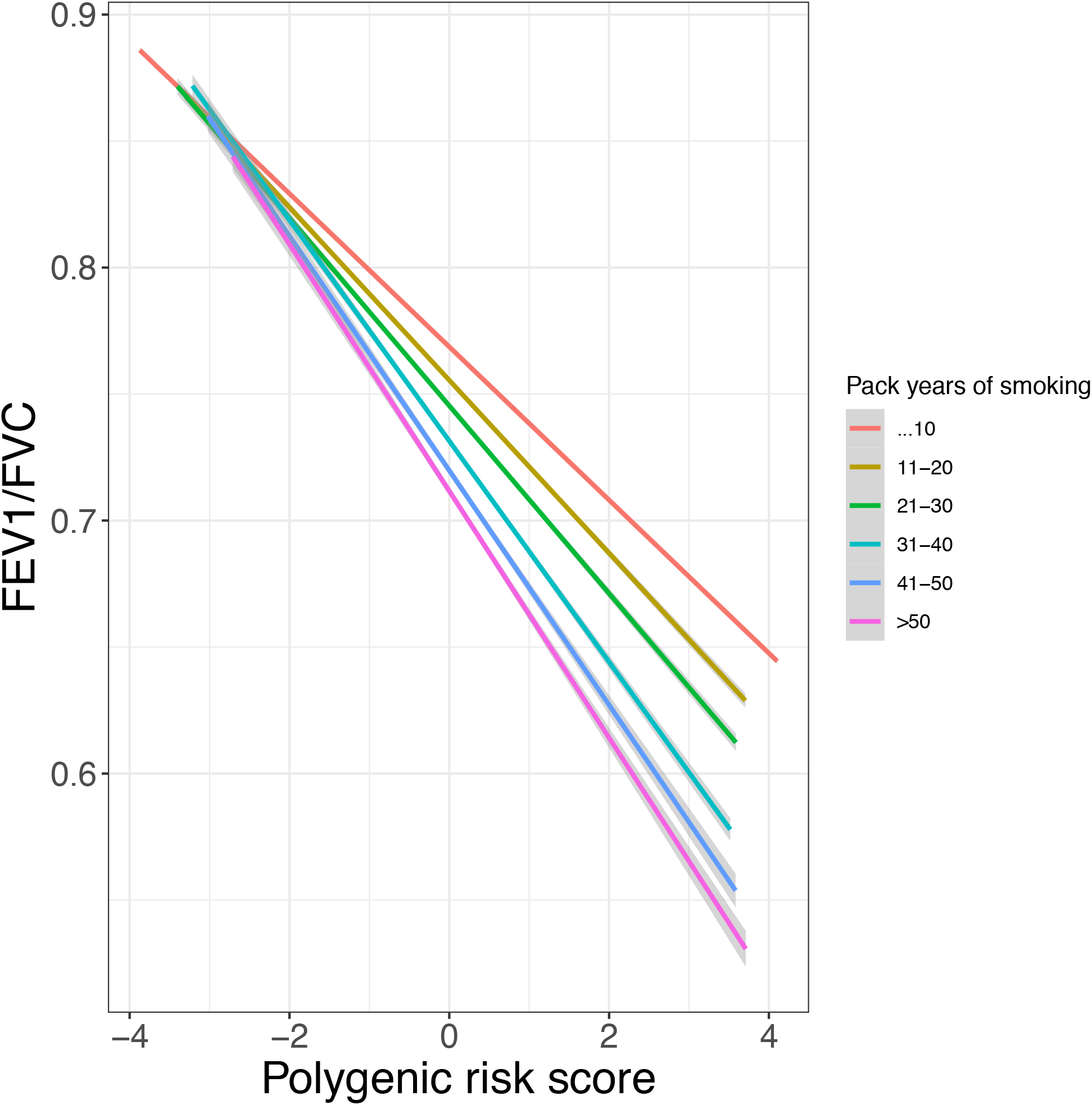
The relationship between PRS and FEV1/FVC by pack years of smoking.

The relationship between the PRS and FEV_1_/FVC stratified by ever-vs. never- and current-vs. former-/never-smoking statuses are shown in Figures S3A and B, respectively. Ever-smoking and the PRS*ever-smoking status interaction term were significantly associated with FEV_1_/FVC (both p < 0.0001, Table S3). Similarly, current smoking status and the PRS*current-smoking status interaction term were significantly associated with FEV_1_/FVC (both p < 0.0001, Table S4). In stratified analyses, we observed similar results between PRS and smoking exposures (Table S5, Figure S4). Additionally, ever smoking status and the PRS*ever-smoking status interaction term were significantly associated with FEV_1_/FVC in the lowest (β [interaction]=-0.0033 [95% CI: −0.0058,-0.00085], p = 0.0082) and highest (β [interaction]=-0.0095 [95% CI: −0.013,-0.0056], p < 0.0001) deciles of predicted genetic risk (Table S5).

The joint effects of pack-years of smoking categories and PRS are shown in Figure 2. Being in the lowest decile of predicted genetic risk and having more than 50 pack-years of smoking exposure (β=-0.022 [95% CI: −0.026, −0.018], p < 0.0001) had a similar estimated effect size as being in the highest decile of genetic risk and having 11-20 pack-years of smoking exposure (β=-0.024 [95% CI: −0.026,-0.023], p < 0.0001). We observed a greater effect size of being in the highest decile of genetic risk and having more than 50 pack-years of smoking exposure (β=-0.051 [95% CI: −0.054,-0.047]) than would be expected, confirming possible interaction between the PRS and pack-years of smoking; we observed a similar association for those in the highest genetic risk decile who were current smokers (Table S6), but a non-significant difference for ever smokers in the highest genetic risk decile (Table S7).

**Figure 2:**
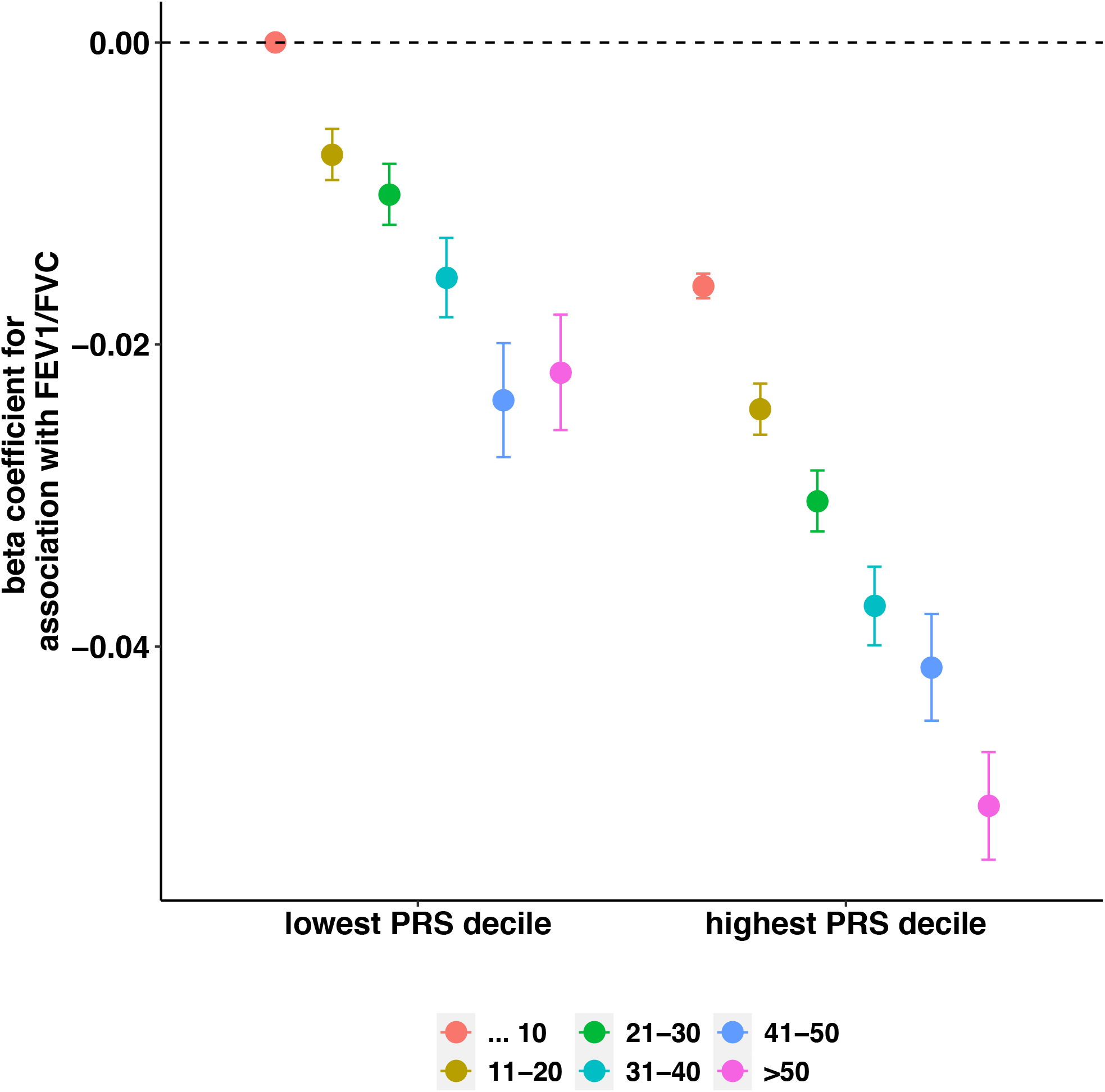
Joint effects of pack years of smoking category and PRS decile. PRS = polygenic risk score.

### Norms of reaction for highest versus lowest predicted genetic risk deciles

In Figure 3, we show different norms of reaction for the effects of pack-years of smoking on FEV_1_/FVC among those in the highest vs. lowest deciles of predicted genetic risk. For any given level of pack-years of smoking, those in the highest PRS decile had lower FEV_1_/FVC compared to those in the lowest decile of PRS. Analysis of covariance confirms the slopes of the lines (Figure 3) are significantly different (p < 0.0001). We observed similar results in ever-smokers (Figure S5). For every 20 pack years of smoking, those in the first decile had a change of - 0.0084 (95% CI: −0.0091, −0.0076) in FEV_1_/FVC, while those in the 10^th^ decile of predicted genetic risk had a change of −0.017 (95% CI: −0.018, −0.016), representing an approximately twofold reduction in FEV_1_/FVC for each 20 pack-years of smoking for those in the highest compared to the lowest decile of predicted genetic risk.

**Figure 3:**
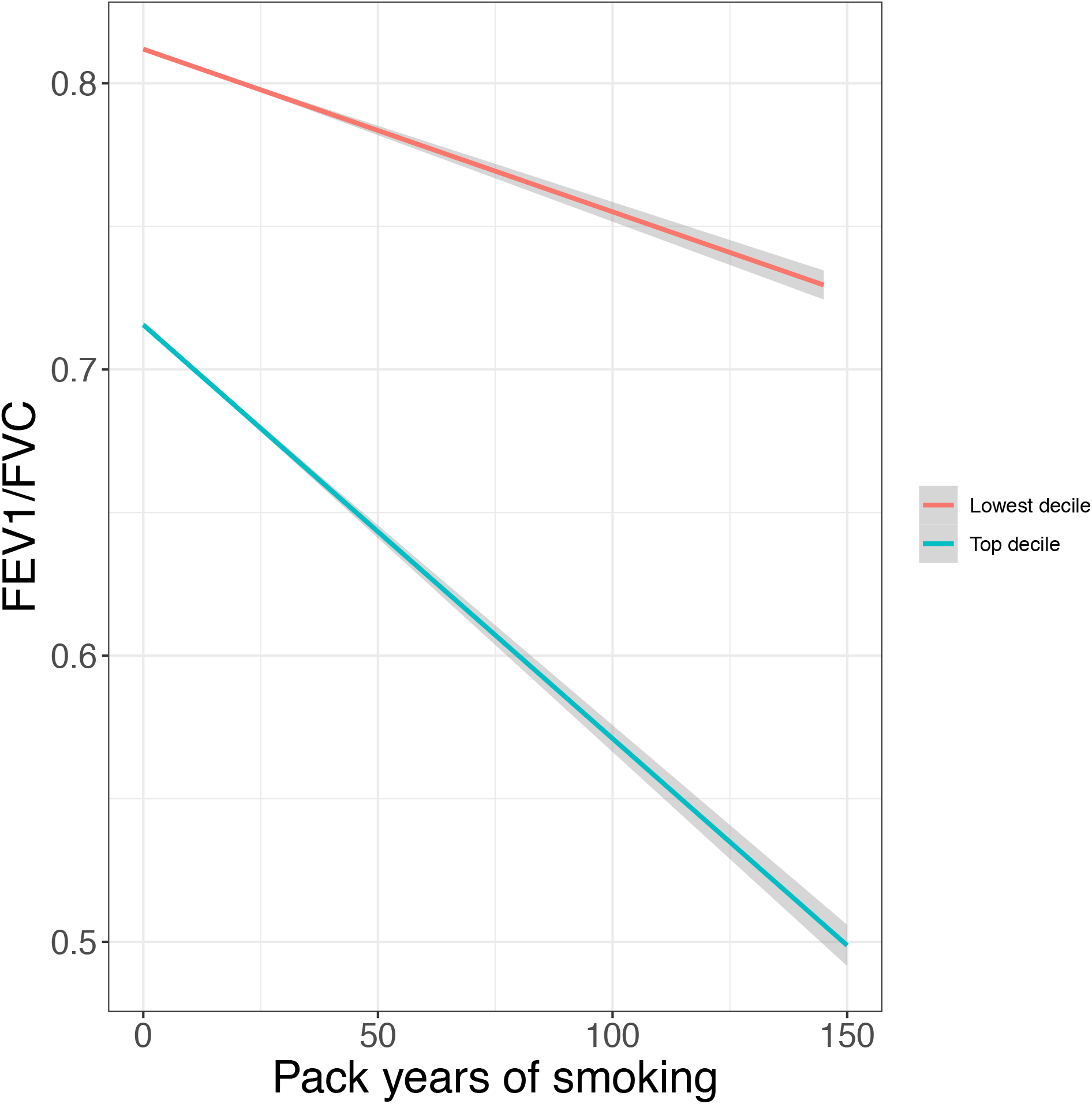
Norms of reaction for those with high versus low predicted genetic risk for COPD. The slopes of these lines were significantly different in analysis of covariance (p < 0.0001).

## Discussion

In this study of over 300,000 UK Biobank participants, we found three measures of smoking exposure modified the effect of a polygenic risk score (PRS) on the quantitative measure of lung function (FEV_1_/FVC). As expected, smoking was detrimental to lung function across all categories of predicted genetic risk. For any given level of pack-years of smoking exposure, however, those at highest genetic risk showed lower FEV_1_/FVC than those with the lowest predicted genetic risk. The effects of heavy smoking and being in the highest decile of predicted genetic risk were greater than would be expected based on the additive effects of both risk factors. These results support the idea that diminished pulmonary function (a measure of airflow obstruction) are, at least partially, due to gene-by-smoking interactions, and those in higher genetic risk categories are more susceptible to the deleterious effects of smoking.

Compared to previous studies, our study included more participants, leveraged a more powerful measure of genetic predisposition for low lung function (i.e. PRS), examined three different measures of smoking exposure (pack-years, ever smoking, current smoking), and examined ‘norms of reactions’ for those in the highest compared to the lowest deciles of predicted genetic risk. Our findings are consistent with Aschard et al.^23^ who reported an interaction between ever-smoking status and a genetic risk score for FEV_1_/FVC based on 26 different variants. In a family-based study of the rs28929474 variant (Z allele) in *SERPINA1*, which leads to alpha-1 antitrypsin deficiency (AATD) and greatly increased risk of emphysema, there was a significant genotype-by-smoking interaction on FEV_1_ ^30^. A strong smoking interaction of rs28929474 heterozygote status with ever-smoking on lung function and COPD has been reported in UK Biobank^31^. Genome-wide interaction studies have confirmed the interaction of smoking with rs28927474 on risk to COPD^11^, and identified gene-by-smoking interactions at several other loci^8–14^.

By contrast, Shrine et al.^19^ constructed a genetic risk score from 279 variants shown to influence lung function, but did not observe any evidence of interaction with ever-smoking status on FEV_1_/FVC; further, the authors reported an interaction with moderate-to-severe COPD status in the opposite direction as expected. A prior GWAS selected ∼50,000 individuals with low, average, and high FEV_1_, reported no gene-smoking interactions at genome-wide significance^32^. The reasons for disparate findings between these studies and the current one are unclear, but may represent the effects of different loci with varying degrees of interaction effect ^4,19,20,33^. While the PRS used in the current study was derived from the Shrine et al. GWAS results^19^, not all variants reached genome-wide significance, and consequently, included many more variants (∼2.5 M); this more refined level of predicted genetic risk may have provided the power to detect gene-by-smoking interactions.

Smoking was detrimental even to those with low predicted genetic risk, and the effects were greater for those with high predicted genetic risk. For any given level of pack-years of smoking, those in the highest decile had lower FEV_1_/FVC compared to those in the lowest decile of predicted genetic risk. These findings are in contrast to observations in cardiovascular disease, where the association between smoking and coronary heart disease was greater for those in the lowest compared to the highest tertile of a PRS^22^. This difference may reflect that many individuals can develop coronary disease in the absence of cigarette smoking, and that smoking is a greater risk factor for those with low polygenic risk for coronary disease. Meanwhile, airflow obstruction primarily occurs in the setting of cigarette smoking exposure. Furthermore, those with low predicted genetic risk and high smoking exposure had similar risk for low FEV_1_/FVC as those with high genetic risk and low smoking exposure. Taken together, these results emphasize that abstaining from smoking is crucial to preventing obstructive lung disease regardless of an individual’s predicted genetic risk, and that those in the highest risk groups might benefit from intensive smoking cessation measures with respect to the phenotypes examined in this study.

Our results suggest that the PRS includes variants that represent biological pathways by which smoking exerts deleterious effects. Some of these variants may act to confer resilience^34,35^ or susceptibility to the effects of cigarette smoke. Further investigation into the role of specific variants in susceptibility to cigarette smoke may be performed by inclusion of other “Omics” data types and leveraging network analytic techniques could help elucidate mechanisms of resilience and susceptibility. For example, the effect of occupational exposures was modified by rs9931086 in *SLC38A8* on FEV_1_, and network analyses suggested inflammatory processes involving *CTLA-4, HDAC*, and *PPAR-alpha*, may provide mechanistic links for the observed interaction^36^; however, this was a small study that needs replication.

Strengths of this study include use of a large volunteer cohort, utilizing the most powerful measure for genetic risk for low lung function available to date (i.e. a genome-wide PRS), and comparing individuals at extremes of predicted genetic risk. Limitations, inherent to study design, include that the UK Biobank is a single cohort observed in cross-section. Examining the effects of gene-by-smoking interactions on incident COPD should be pursued. We were not able to model the time-varying effects of smoking exposure. The PRS was partially developed using samples from UK Biobank, leading to overfitting of the PRS with respect to spirometric measures; while this issue should not affect interaction assessments, these results should ideally be replicated in future studies. However, the strong effect sizes and robustness to stratified and transformed analyses does lend confidence to our results. We included European-ancestry participants only because the PRS was derived solely from Europeans. Identification of causal variants and genetic prediction in single ancestry populations demonstrate limited portability to multi-ancestry populations^37,38^. The 279 lung function variants from Shrine et al.^19^ was curated to ensure variants for smoking behavior were excluded, but the PRS used in the current study included ∼2.5 million variants and was not similarly curated. Including variants that are causal for smoking behavior could bias the interaction term^39^; however, there was a very weak correlation between this PRS and smoking exposure in UK Biobank, and previously no correlation with smoking in case-control cohorts was observed^20^, suggesting that the PRS used in the current study largely reflects the genetics of lung function. Finally, we emphasize that smoking cessation is the main preventive intervention to airflow obstruction regardless of genetic susceptibility. Our interaction findings further support a more intensive smoking cessation is required to those with high genetic susceptibility to diminished lung function. The effectiveness of targeted genetically-informed smoking cessation interventions is unclear, though there is evidence that knowledge of genetic risk for AATD can increase smoking cessation^40^. Clinical utility of the PRS will depend on dissecting biological mechanisms of susceptibility to the harmful effects of smoking.

In conclusion, diminished FEV_1_/FVC and airflow obstruction, which are characteristic of COPD, may be partially attributable to gene-by-smoking interactions. As expected, smoking was harmful across all genetic risk groups, but worse for those in the highest decile of predicted genetic risk. Large-scale replication and further investigations into mechanisms of interaction are needed.

## Supporting information

Suppplementary

## Data Availability

None.

## Author Contributions

Study Design: Matthew Moll, Woori Kim, Terri H. Beaty, Michael H. Cho

Acquisition, analysis, or interpretation of the data: Woori Kim, Matthew Moll, Brian D. Hobbs, Michael H. Cho, Edwin K. Silverman, Terri H. Beaty.

Critical revision of the manuscript for important intellectual content: All authors

Statistical analysis: Matthew Moll, Woori Kim, Brian D. Hobbs, Michael H. Cho, Dandi Qiao, Edwin K. Silverman

Obtained funding: Terri H. Beaty, Edwin K. Silverman, Michael H. Cho.

