## Supplementary material for "Smoking Interaction with a Polygenic Risk Score for Reduced Lung Function": Suppplementary

**Supplementary Note**

Table of Contents

Supplementary Tables

Table S1…………………………………………………………………………...……2

Table S2………………………………………………………………………………...3

Table S3………………………………………………………………………………...4

Table S4………………………………………………………………………………...5

Table S5………………………………………………………………………………...6

Table S6………………………………………………………………………………....7

Table S7………………………………………………………………………………....8

Supplementary Figures

Figure S1……………………………………………………………………………….9

Figure S2……………………………………………………………………………….10

Figure S3……………………………………………………………………………….11

Figure S4……………………………………………………………………………….13

Figure S5……………………………………………………………………………….15

References………………………………………………………………………………………16

**Supplementary Tables**

Table S1: Multivariable linear regression of the form FEV_1_/FVC ~ PRS + Pack years of smoking + PRS*pack-years of smoking + covariates. Covariates include age, age^2^, sex, height, genotyping array, principal components of genetic ancestry. Pack years of smoking is included as a continuous variable. PRS = polygenic risk score ^1,2^.

| *Variable* | *Beta (95% CI)* | *P* |
| --- | --- | --- |
| PRS | -0.03 (-0.031--0.03) | < 1e-256 |
| Pack years | -0.0064 (-0.0066--0.0063) | < 1e-256 |
| PRS X Pack years | -0.0028 (-0.0029--0.0026) | < 1e-256 |

Table S2: Multivariable linear regression of the form FEV_1_/FVC ~ PRS + Pack years of smoking + PRS*pack-years of smoking + covariates. Covariates include age, age^2^, sex, height, genotyping array, principal components of genetic ancestry. Pack years of smoking were transformed prior to analyses to ensure robustness of results to any misspecification of the main effects of smoking. PRS = polygenic risk score.

| **Variable** | **Beta (95% CI)** | **P** |
| --- | --- | --- |
| *Log-transformed pack years* | | |
| PRS | -0.021 (-0.022--0.022) | 1.30E-204 |
| Pack years | -0.0078 (-0.0083--0.0083) | 1.60E-231 |
| PRS X Pack years | -0.0058 (-0.0063--0.0063) | 4.50E-138 |
| *Scaled and centered pack years* | | |
| PRS | -0.033 (-0.034--0.034) | <1e-256 |
| Pack years | -0.0072 (-0.0076--0.0076) | <1e-256 |
| PRS X Pack years | -0.004 (-0.0044--0.0044) | 1.30E-124 |
| *Rank-normalized pack years* | | |
| PRS | -0.027 (-0.028--0.028) | <1e-256 |
| Pack years | -0.016 (-0.017--0.017) | <1e-256 |
| PRS X Pack years | -0.0095 (-0.01--0.01) | 1.10E-141 |

Table S3: Multivariable linear regression of the form FEV_1_/FVC ~ PRS + Ever smoking status + PRS*Ever smoking status + covariates. Covariates include age, age^2^, sex, height, pack years of cigarette smoking, genotyping array, principal components of genetic ancestry. Ever-smoking status was compared to never-smoking status. PRS = polygenic risk score.

| *Variable* | *Beta (95% CI)* | *P* |
| --- | --- | --- |
| PRS | -0.03 (-0.031--0.03) | < 1e-256 |
| Ever smoker | -0.016 (-0.016--0.015) | < 1e-256 |
| PRS X Ever smoker | -0.0064 (-0.0068--0.006) | 6.00E-209 |

Table S4: Multivariable linear regression of the form FEV_1_/FVC ~ PRS + Current smoking status + PRS*Current smoking status + covariates. Covariates include age, age^2^, sex, height, pack years of cigarette smoking, genotyping array, principal components of genetic ancestry. Current smoking status was compared to former-/never smoking status. PRS = polygenic risk score.

| *Variable* | *Beta (95% CI)* | *P* |
| --- | --- | --- |
| PRS | -0.032 (-0.032--0.032) | < 1e-256 |
| Current smoker | -0.027 (-0.028--0.027) | < 1e-256 |
| PRS X Current smoker | -0.0091 (-0.0097--0.0084) | 5.60E-157 |

Table S5: The cohort was stratified by ever- versus never-smoking status and highest versus lowest PRS decile. Interaction terms were evaluated between dichotomized PRS (denoted "Top decile of PRS") or continuous PRS (denoted "PRS") and the appropriate smoking variable. Each box represents a single model. PRS = Polygenic risk score.

| *Stratum* | *Variable* | *Beta (95% CI)* | *P* |
| --- | --- | --- | --- |
|  | Ever smoking | -0.014 (-0.018--0.01) | 6.00E-13 |
| Lowest PRS decile | PRS | -0.024 (-0.025--0.022) | 1.30E-164 |
|  | PRS X Ever smoking interaction | -0.0033 (-0.0058--0.00085) | 0.0082 |
|  | Ever smoking | -0.011 (-0.017--0.0042) | 0.0013 |
| Highest PRS decile | PRS | -0.037 (-0.04--0.035) | 1.30E-159 |
|  | PRS X Ever smoking interaction | -0.0095 (-0.013--0.0056) | 2.10E-06 |
| Never smokers | PRS | -0.03 (-0.03--0.03) | < 1e-256 |
| Never smokers | Top decile of PRS | -0.095 (-0.097--0.094) | < 1e-256 |
|  | Pack years of cigarette smoking | -0.00047 (-0.00049--0.00045) | < 1e-256 |
| Ever smokers | PRS | -0.031 (-0.032--0.031) | < 1e-256 |
|  | PRS X Pack years of smoking interaction | -0.00026 (-0.00028--0.00024) | 1.20E-134 |
|  | Pack years of cigarette smoking | -3.1e-05 (-0.00011-4.6e-05) | 0.43 |
| Ever smokers | Top decile of PRS | -0.1 (-0.1--0.099) | < 1e-256 |
|  | Top decile of PRS X Pack years of smoking interaction | -0.00075 (-0.00084--0.00065) | 3.50E-53 |

Table S6: Joint effects of current-smoking status and highest versus lowest decile of PRS. PRS = polygenic risk score.

|  | *Lowest decile of PRS (beta (95% CI))* | *Highest decile of PRS (beta (95% CI))* |
| --- | --- | --- |
| Former-/Never-smokers | 0 (ref) | -0.017 (-0.017--0.016) |
| Current smokers | -0.016 (-0.018--0.013) | -0.037 (-0.039--0.035) |

Table S7: Joint effects of ever-smoking and being in the highest vs. lowest decile of the PRS. PRS = polygenic risk score.

|  | *Lowest decile of PRS (beta (95% CI))* | *Highest decile of PRS (beta (95% CI))* |
| --- | --- | --- |
| Never smokers | 0 (ref) | -0.016 (-0.017--0.015) |
| Ever smokers | -0.0026 (-0.0038--0.0013) | -0.022 (-0.023--0.02) |

**Supplementary Figures**

Figure S1: Participants included in study.

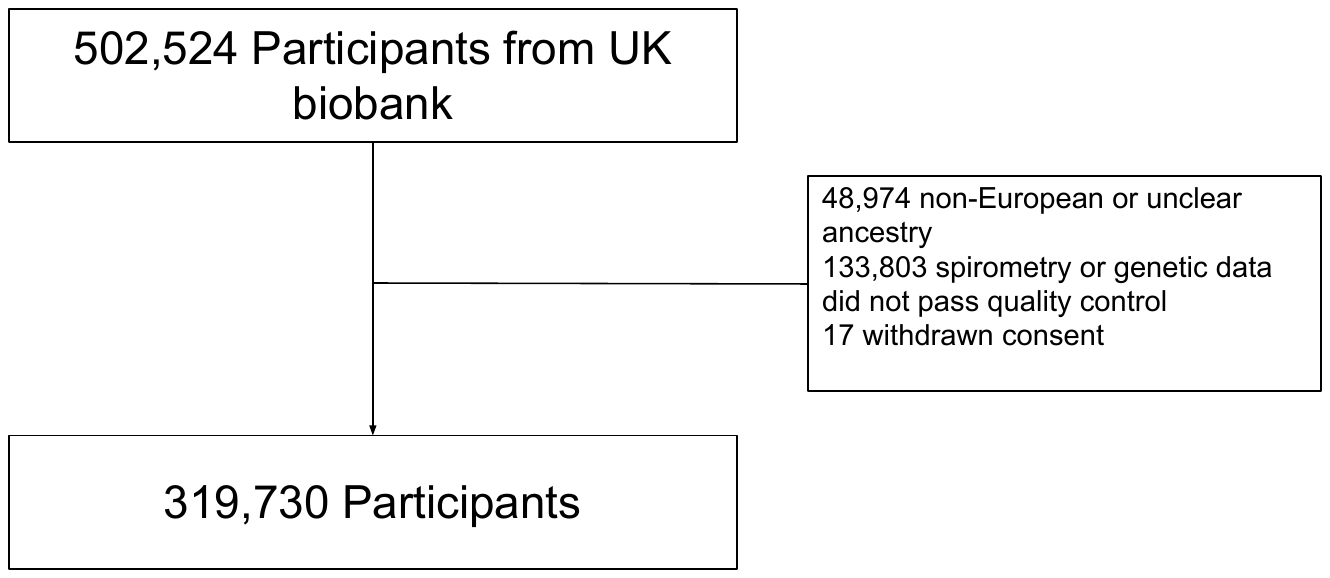

Figure S2: Scatterplot of the polygenic risk score and pack-years of smoking exposure (Pearson r-value = 0.041, p < 0.0001).

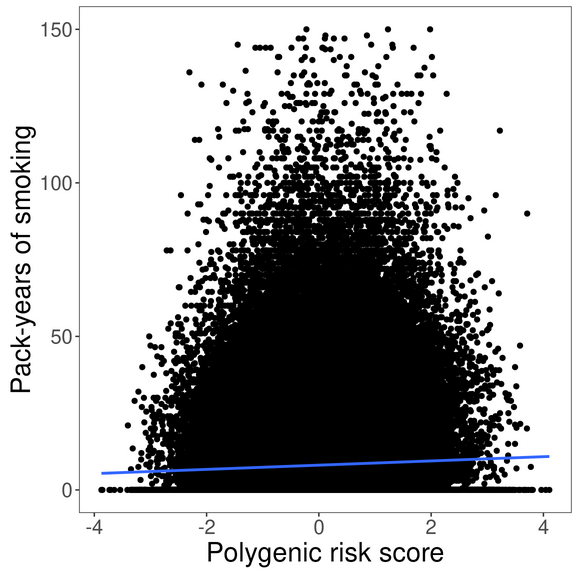

Figure S3: The relationship between the polygenic risk score and FEV_1_/FVC in ever- vs. never-smokers (A) and current- vs. former-/never- (B) smokers.

A)

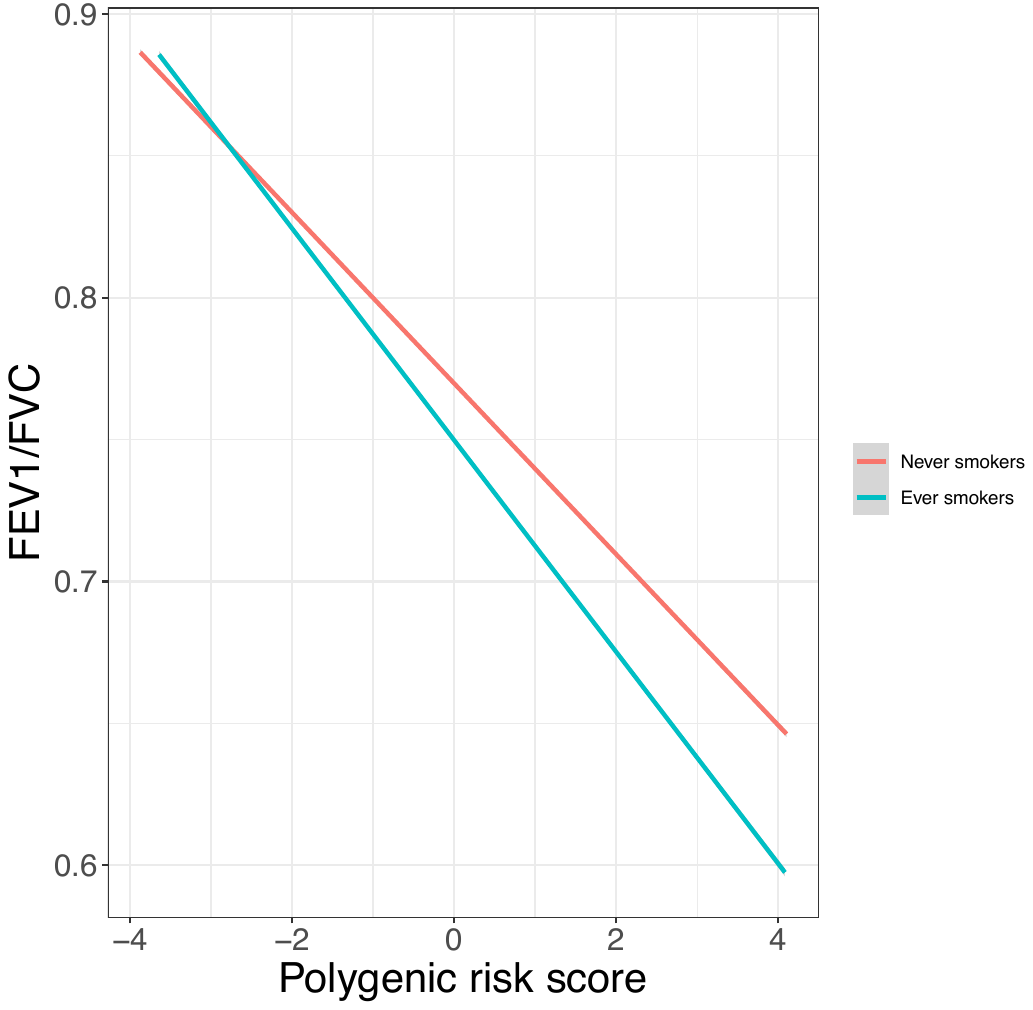

B)

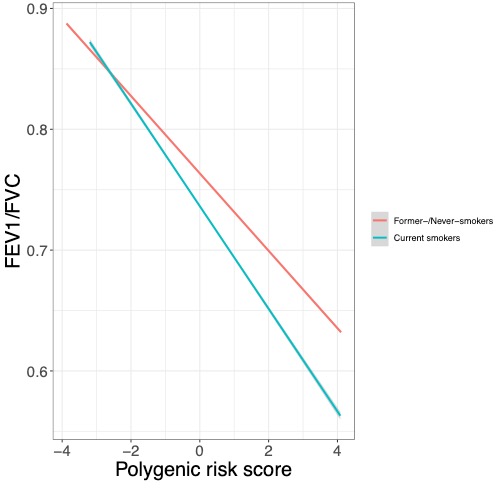

Figure S4: The relationship between PRS and FEV_1_/FVC by pack years of smoking (A) and current versus former smoking status (B) in ever smokers (n=146,679). Interaction terms and p-values are shown where appropriate.

A)

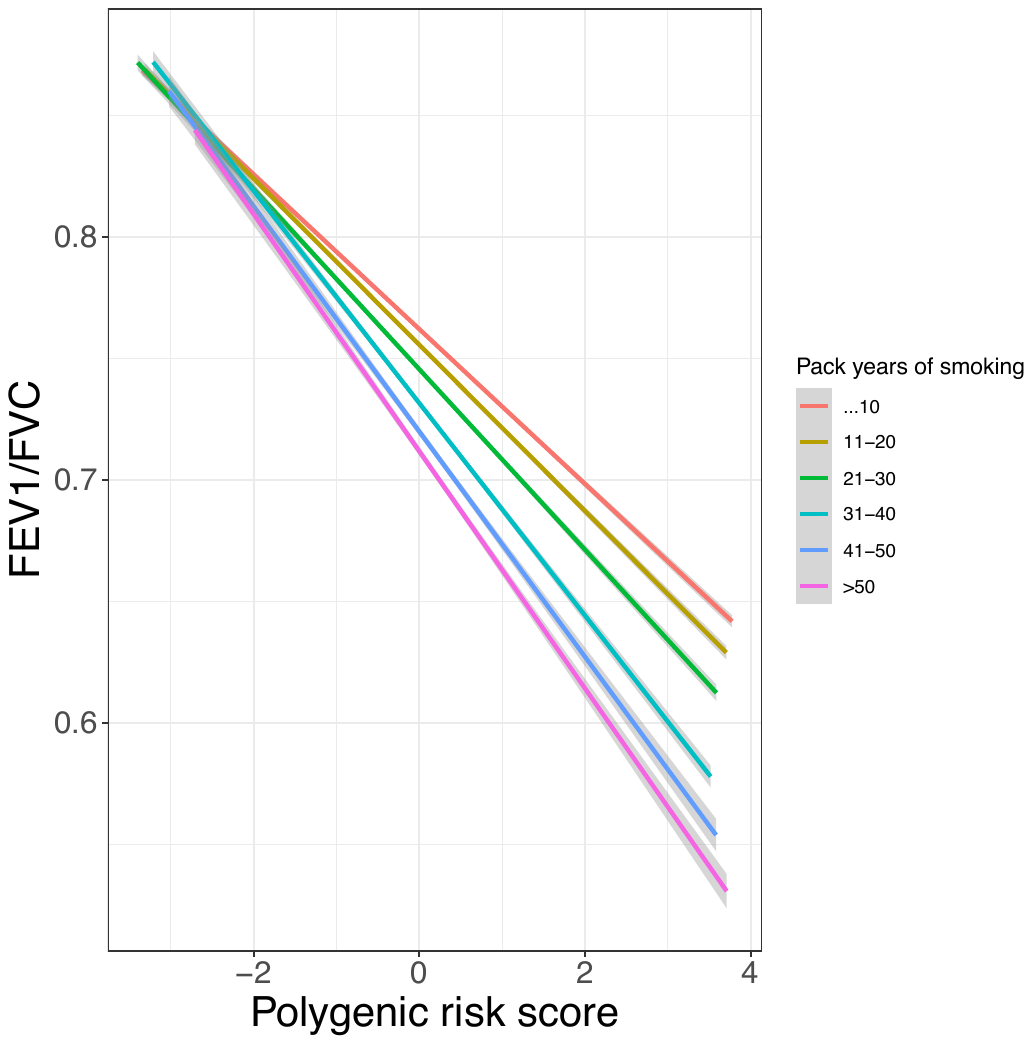

B)

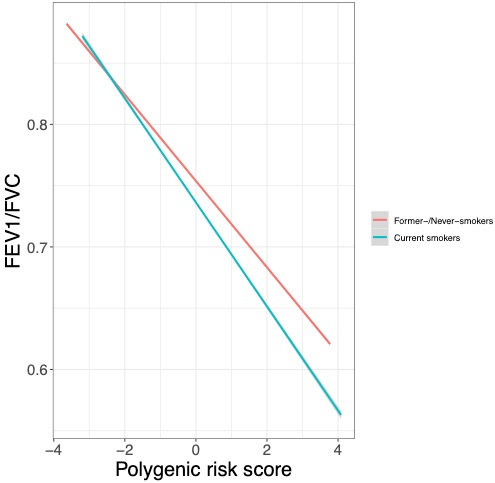

Figure S5: Norms of reaction for those with high versus low genetic risk for COPD in ever smokers (n=146,679). The slopes of these lines were significantly different in analysis of covariance (p < 0.0001).

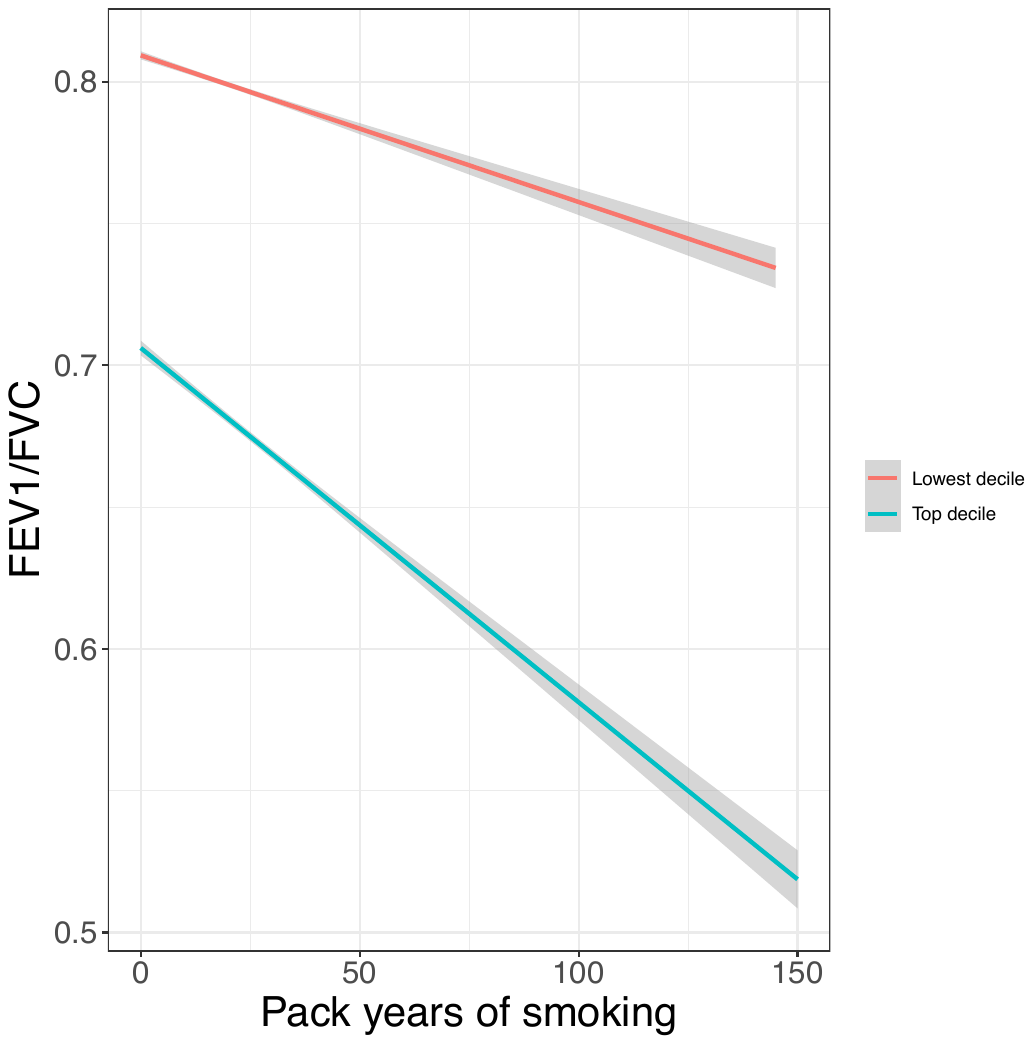
